# Integrated Clinical, Climate, and Environmental Prediction Modeling for Diagnosis of Spotted Fever Group Rickettsioses in northern Tanzania

**DOI:** 10.1101/2024.06.20.24309257

**Authors:** Robert J. Williams, Ben J. Brintz, William L. Nicholson, John A. Crump, Ganga Moorthy, Venace P. Maro, Grace D. Kinabo, James Ngocho, Wilbrod Saganda, Daniel T. Leung, Matthew P. Rubach

## Abstract

Spotted fever group rickettsioses (SFGR) pose a global threat as emerging zoonotic infectious diseases; however, timely and cost-effective diagnostic tools are currently limited. While traditional clinical prediction models focus on individual patient-level parameters, we hypothesize that for infectious diseases, the inclusion of location-specific parameters such as climate data may improve predictive ability. To create a prediction model, we used data from 449 patients presenting to two hospitals in northern Tanzania between 2007 to 2008, of which 71 (15.8%) met criteria for acute SFGR based on ≥4-fold rise in antibody titers between acute and convalescent serum samples. We fit random forest classifiers by incorporating clinical and demographic data from hospitalized febrile participants as well as satellite-derived climate predictors from the Kilimanjaro Region. In cross-validation, a prediction model combining clinical, climate, and environmental predictors (20 predictors total) achieved a statistically non-significant increase in the area under the receiver operating characteristic curve (AUC) compared to clinical predictors alone [AUC: 0.72 (95% CI:0.57-0.86) versus AUC: 0.64 (95% CI:0.48-0.80)]. In conclusion, we derived and internally-validated a diagnostic prediction model for acute SFGR, demonstrating that the inclusion of climate variables alongside clinical variables improved model performance, though this difference was not statistically significant. Novel strategies are needed to improve the diagnosis of acute SFGR, including the identification of diagnostic biomarkers that could enhance clinical prediction models.

## Background

Spotted fever group rickettsioses (SFGR) are a group of illnesses caused by bacteria from the genus *Rickettsia* that include endemic, new, and emerging zoonotic infectious diseases with a worldwide distribution. In several African countries, SFGR has been identified as the infectious etiology in 5-22% of febrile hospital admissions.^1–3^ Prompt recognition and treatment of SFGR are important as multiple studies have shown the delay in initiation of tetracycline antimicrobials is associated with increased morbidity and mortality.^4–6^ However, current diagnostic methods do not allow for timely and accurate diagnosis. The most sensitive reference standard diagnostic, a ≥4-fold rise in immunofluorescent antibody (IFA) titer between paired acute and convalescent serum samples, requires convalescent serum collection, and therefore by definition cannot establish the diagnosis at the time of presentation.^7^ In the case of *R. africae* and *R. conorii,* SFGR of importance in African countries, seroconversion may not occur until 4 weeks after illness onset.^8^

*Rickettsia* are intracellular species and do not circulate extensively in the bloodstream, limiting the sensitivity of polymerase chain reaction (PCR) on blood specimens to around 60%.^7^ Laboratory values such as thrombocytopenia, hyponatremia, and elevated transaminases are supportive features, but cannot be relied on to guide early management as they are non-specific findings and often within normal limits or only slightly above the reference range early in the course of illness.^9^ Clinical diagnosis relying on the triad classically associated with SFGR—a history of a tick bite, rash, and fever—only occur in a minority of cases.^9–12^ Incorporating tetracycline therapy into the empiric syndromic management of febrile illness in high prevalence settings would not only exacerbate antimicrobial resistance, but subject children to the risks of tetracycline therapy, including bone growth suppression and permanent tooth discoloration.^13,14^ Thus, more accurate, timely, and cost-effective tools are needed for diagnosis of SFGR.

Clinical Decision-Support Systems (CDSS) incorporating prediction models have the potential to improve management of infectious diseases. CDSS have proven effective at enhancing therapeutic management and reducing unnecessary diagnostic tests in both high-income countries (HICs) ^15^ and LMICs. ^16–18^ Traditional predictive models generally incorporate clinical information that is obtained solely from the presenting patient. However, as with other zoonoses, the range and incidence of SFGR has been associated with climate and climate-related environmental factors.^19,20^ Thus, incorporating location-specific parameters, such as climate and environmental data, into a prediction model may increase diagnostic accuracy.^18,21,22^

In this study, we demonstrate a ‘proof of concept’ by integrating location-specific parameters into clinical prediction. Our overarching goal is to create an accessible and cost-effective CDSS that assists clinicians in diagnosing SFGR. To achieve this, we used data from a clinical study of febrile illness in an SFGR-endemic region in northern Tanzania^23^ to develop a clinical prediction model that incorporates climate and environmental data.

## Methods

### Study Design, Setting, and Data Source

For the derivation and validation of a prediction model, we used de-identified data from a study of participants presenting with febrile illness to two hospitals in Kilimanjaro Region of northern Tanzania, 2007-2008.^24,25^

Prior to data collection, the Kilimanjaro Region in northern Tanzania had a population of 1,380,000 in a mostly rural and semirural setting;^26,27^ and Moshi, the administrative center of the Kilimanjaro Region, had a population of approximately 144,000. ^27^ The climate is characterized by a long rainy period (March-May) and a short rainy period (November-December). Febrile participants presenting to Kilimanjaro Christian Medical Centre (KCMC) or Mawenzi Regional Hospital (MRH) in Moshi, Tanzania from September 2007 through August 2008 were eligible for enrollment.

Complete study methods have been described elsewhere.^24,25^ KCMC is a tertiary care hospital serving several Regions in northern Tanzania; at the time of the study KCMC had 458 inpatient beds. MRH, the regional hospital for Kilimanjaro, had 300 beds at the time of the study and served. Together, KCMC and MRH served as major providers of hospital-based care in the Moshi area.

For pediatric participants 2 months to 12 years, inclusion criteria were a history of fever in the past 48 hours or a measured axillary temperature ≥ 37.5°C or rectal temperature ≥ 38°C. For adolescent and adult participants (≥13 years of age), inclusion criteria were an oral temperature ≥38°C on admission to the hospital. All participants required paired sera for inclusion in this analysis. Blood specimens were collected for a complete blood count (CBC) and serologic infectious disease diagnostics. Participants were also tested for HIV and malaria with rapid diagnostic testing. After obtaining informed consent, a trained study team member collected standardized demographic data, clinical history, and physical examination findings. Participants were asked to return 4-6 weeks after enrollment for collection of a convalescent serum sample.

Acute and convalescent serum samples collected for SFGR testing were sent to the Rickettsial Zoonoses Branch of the US Centers for Disease Control and Prevention (US CDC). Serum samples were tested for SFGR by IgG IFA to *R. conorii* (Moroccan strain). SFGR was defined as a ≥4- fold increase in IFA titer to *R. conorii* between acute and convalescent serum in a participant. Participants with less than a 4-fold rise in IFA titer to *R. conorii* were considered non-SFGR febrile illness.

For each participant, a trained study team member collected standardized demographic data, clinical history, admission vital signs, and physical examination findings. As the presentation of infectious disease can differ between pediatric and adult participants, several of the clinical variables were only obtained for either pediatric or adult participants. For our clinical prediction modeling we only included variables that were collected for both groups, which included: age, heart rate, respiration rate, blood pressure, oxygen saturation, height, weight, body mass index (BMI), cough, diarrhea, emesis, hematochezia, dyspnea, seizures, crepitations, hepatomegaly, splenomegaly, pallor, lymphadenopathy, oral candidiasis, meningeal signs, HIV rapid diagnostic result, malaria rapid diagnostic result, and if the participant resided in a rural setting. We recorded clinical symptoms and physical exam findings as binary variables and age, vital signs, and complete blood count (CBC) results as continuous variables.

### Climate and Environmental data

We extracted climate and environmental data from the MODIS (Moderate Resolution Imaging Spectroradiometer) satellite for the Kilimanjaro Region, Tanzania. Environmental data included the normalized difference vegetation index (NDVI), the enhanced vegetation index (EVI), and the normalized difference water index (NDWI), evapotranspiration. Climate data included daytime and nighttime temperature. NDVI and EVI indicators are based on a 16-day time series composite image at 1km * 1km spatial resolution and were obtained from Moderate Resolution Imaging Spectroradiometer (MODIS) product MOD13A2. Surface temperatures, acquired from MODIS product MOD11A1, are daily measurements at 1km * 1km spatial resolution. Evapotranspiration is based on an 8-day time series composite image at 500m x 500m spatial resolution and obtained from MOD16A2GF. Finally, using MODI09A1, a 500m x 500m 8-day composite time series, we calculated NDWI from near-infrared (NIR – MODIS band 2) and short- wave infrared (SWIR- MODIS band 6) reflectance’s.^28,29^ We consolidated all climate and environmental data within a uniform 16-day time series window; for shorter times-series, we computed the mean for each 16-day window. For example, data from January 1 through January 16, 2007 constituted one window, followed by measurements from January 17 through February 2, 2007 in the subsequent window. To account for recent climate and environmental patterns that may influence SFGR incidence, we lagged each 16-day times series at one, two, and three months. Finally, we aligned the lagged measurements with the admission dates of study participants. For instance, a participant presenting on April 4, 2007 would have data from the window containing March 4 (1-month lag), February 4 (2-month lag), and January 4 (3-month lag).

### Statistical Analysis and Modeling

To compare acute SFGR versus non-SFGR febrile illness groups on univariate analysis, we used the Wilcoxon rank sum test due to the non-normal distribution of age data. We used Pearson’s Chi-squared test and Fisher’s exact test for categorical variables. We used the random forest algorithm to fit a model to predict risk of participants having acute SFGR versus non-SFGR febrile illness. Random forests are a machine learning algorithm which constructs a multitude of decision trees and averages over them to obtain a prediction robust to nonlinearities and interactions between covariates; random forests algorithms have been widely applied to biomedical sciences for both classification and regression.^30,31^

We excluded predictors with highly skewed binary predictors (predictors with 95% or more values concentrated as either 0 or 1) from analysis. For the remaining predictors, we imputed missing data using the ‘missRanger’ package in R. To determine which predictors to include in our analysis, we fit two distinct models – one utilizing solely satellite-derived climate and environmental data and another incorporating clinical and demographic data. We used the ‘permimp’ package in R to assess the variable importance using permutation-based methods. This method involves systematically shuffling or permuting the values of individual predictors to evaluate their impact on performance. Next, we identified the top 10 predictors from each model based on their respective permuted importance scores. We included these predictors in our final analysis.

To assess predictive performance for each random forest model, we used repeated cross-validation using 80% training/20% testing splits with 100 iterations. In each iteration, we trained models on 80% of the data, made predictions on the 20% test set, and obtained measures of performance.

We determined overall model performance by averaging the area under the receiver operating characteristic curve (AUC) and confidence intervals across the 100 iterations. To determine statistical significance in the AUC between models we used a bootstrap method over 100 iterations, which involves resampling the data with replacement multiple times, creating bootstrap samples. For each bootstrap sample, we generated receiver operating characteristic (ROC) and computed the difference in AUC between the curves. We completed all analyses using R version 4.2.0, and model development/validation was completed in accordance with the Transparent reporting of a multivariable prediction model for individual prognosis or diagnosis (TRIPOD) checklist (Supplement Table 1).

### Research Ethics

The primary study was approved by the Kilimanjaro Christian Medical University College Health Research Ethics Committee, the Tanzania National Institutes for Medical Research National Health Research Ethics Coordinating Committee, and Institutional Review Boards of Duke University Medical Center, and the US CDC. The secondary data analysis was reviewed by the Institutional Review Board of the University of Utah and determined to be exempt (IRB_00164810). All minors had written informed consent given from a parent or guardian, and all adult participants provided their own written informed consent.

## Results

Of the 870 participants enrolled in the study, 449 (51.6%) underwent follow-up for the collection of convalescent serum. Of these 449 participants, 71 (15.8%) met criteria for acute SFGR (Figure 1).

**Figure 1.**
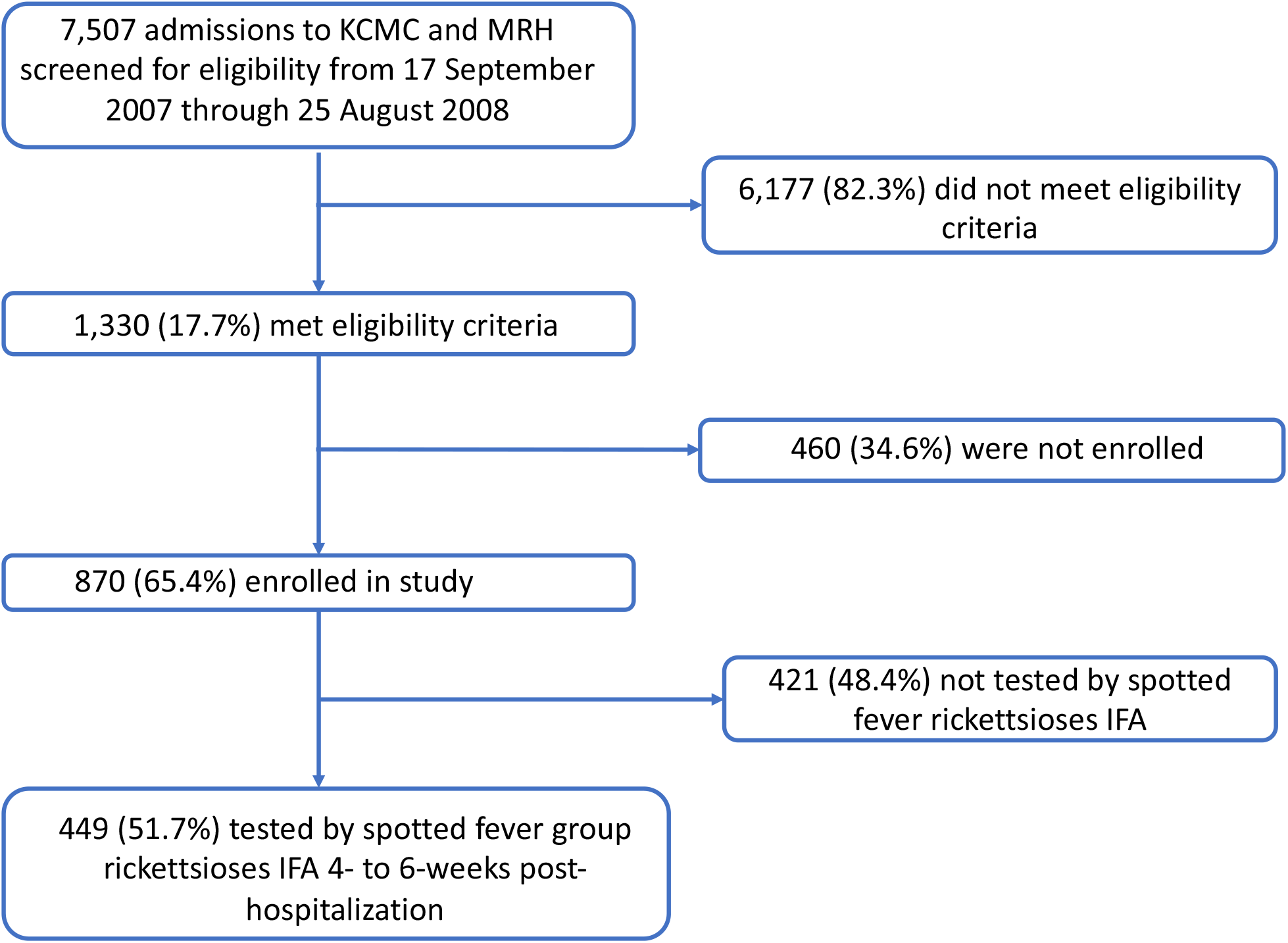
Study flow diagram. Screening and enrollment of patients hospitalized at KCMC and MRH. KCMC: Kilimanjaro Christian Medical Centre; MRH: Mawenzi Regional Hospital; IFA: immunofluorescence assay

We excluded the highly skewed predictors hematochezia, meningeal signs, and the malaria rapid diagnostic from our analysis. We found statistically significant differences in several clinical variables including vital signs, clinical symptoms, and laboratory results between acute SFGR and non- SGFR febrile illness (Table 1). Overall, acute SFGR participants were older (median age 24 versus 8 years, p-value=0.003) with significantly higher height and weight. Acute SFGR participants had a lower respiratory rate than non-SFGR febrile illness participants (median 28 versus 32 breaths per minute, p-value<0.001) and were more likely reside in a rural setting (59% versus 45%, p-value=0.025). There were no significant differences in the CBC results between the two groups, although platelet count and hematocrit had a p-value of 0.07 and 0.08, respectively.

**Table 1.** Clinical characteristics and complete blood count results for febrile participants with and without spotted-fever group rickettsioses, northern Tanzania, 2007-2008.

|  | Overall,<br>N=449<sup>1</sup> | Non-SFGR febrile<br>illness,<br>N=378<sup>1</sup> | Acute SFGR,<br>N=71<sup>1</sup> | p-value<sup>2</sup> |
| --- | --- | --- | --- | --- |
| **Age, years** | 9 (2, 32) | 8 (1, 31) | 24 (4, 39) | 0.003 |
| **Rural** | 211 (47%) | 168 (45%) | 42 (59%) | 0.03 |
| **Admission temperature, °C** | 38.40 (38.00, 39.10) | 38.40 (38.00, 39.00) | 38.60 (38.00, 39.20) | 0.4 |
| **Heart rate, beats per minute** | 112 (97, 131) | 112 (98, 132) | 108 (96, 124) | 0.2 |
| **Respiration rate, breaths per minute** | 32 (24, 44) | 32 (26, 45) | 28 (24, 35) | <0.001 |
| **Oxygen saturation, %** | 96.0 (94.0, 98.0) | 96.0 (94.0, 98.0) | 97.0 (94.0, 98.0) | 0.5 |
| **Systolic blood pressure, mmHg** | 108 (100, 120) | 108 (100, 120) | 108 (100, 117) | 0.4 |
| **Diastolic blood pressure, mmHg** | 68 (60, 74) | 68 (60, 74) | 67 (60, 73) | 0.7 |
| **BMI** | 17.2 (14.4, 21.6) | 17.0 (14.3, 21.0) | 19.2 (15.6, 22.5) | 0.06 |
| **Weight, kg** | 22 (10, 55) | 20 (10, 55) | 49 (14, 61) | 0.01 |
| **Height, m** | 1.25 (0.82, 1.62) | 1.19 (0.81, 1.62) | 1.54 (1.05, 1.63) | 0.01 |
| **Cough** | 293 (65%) | 252 (67%) | 41 (58%) | 0.1 |
| **Oral Candida** | 43 (9.6%) | 39 (10%) | 4 (5.6%) | 0.2 |
| **Crepitations** | 196 (44%) | 165 (44%) | 31 (44%) | >0.9 |
| **Hepatomegaly** | 34 (7.6%) | 27 (7.2%) | 7 (9.9%) | 0.4 |
| **Diarrhea** | 90 (20%) | 77 (20%) | 13 (18%) | 0.7 |
| **Emesis** | 132 (29%) | 109 (29%) | 23 (32%) | 0.5 |
| **Dyspnea** | 152 (34%) | 132 (35%) | 20 (28%) | 0.3 |
| **Seizure** | 41 (9.2%) | 37 (9.8%) | 4 (5.6%) | 0.3 |
| **Pallor** | 32 (7.2%) | 27 (7.2%) | 5 (7.0%) | >0.9 |
| **Lymphadenopathy** | 39 (8.7%) | 34 (9.0%) | 5 (7.0%) | 0.6 |
| **Rapid Malaria Diagnostic** | 19 (4.2%) | 16 (4.2%) | 3 (4.2%) | >0.9 |
| **Rapid HIV Diagnostic** |  |  |  | 0.13 |
| **Positive** | 103 (23%) | 92 (24%) | 11 (15%) |  |
| **Indeterminant** | 8 (1.8%) | 8 (2.1%) | 0 (0%) |  |
| **White blood cell count, K/μL** | 9 (6, 13) | 9 (6, 13) | 7 (5, 13) | 0.1 |
| **Red blood cell count, M/μL** | 4.28 (3.66, 4.79) | 4.24 (3.59, 4.79) | 4.43 (3.97, 4.82) | 0.2 |
| **Hemoglobin, g/dL** | 10.80 (9.00, 12.40) | 10.70 (9.00, 12.28) | 11.30 (9.95, 12.75) | 0.1 |
| **Hematocrit, %** | 32 (28, 37) | 32 (27, 36) | 33 (30, 37) | 0.08 |
| **Platelet count, K/μL** | 267 (161, 391) | 275 (164, 404) | 225 (130, 360) | 0.07 |
| **Neutrophil, %** | 62 (42, 76) | 62 (42, 76) | 65 (45, 79) | 0.3 |
| **Lymphocyte, %** | 25 (16, 43) | 26 (16, 44) | 22 (13, 42) | 0.2 |
| **Monocyte, %** | 8.5 (5.9, 11.5) | 8.5 (5.9, 11.6) | 7.8 (5.3, 10.5) | 0.3 |
| **Eosinophil, %** | 0.20 (0.00, 0.90) | 0.20 (0.00, 1.00) | 0.20 (0.00, 0.70) | 0.3 |
| **Basophil, %** | 0.80 (0.40, 1.20) | 0.80 (0.40, 1.20) | 0.80 (0.45, 1.10) | >0.9 |
| <sup>1</sup> n (%); Median (IQR) |  |  |  |  |
| <sup>2</sup> Pearson's Chi-squared test; Wilcoxon rank sum test; Fisher's exact test |  |  |  |  |

We also found several significantly different climate and environmental predictors between acute SFGR and non-SFGR febrile illness participants (Supplemental table S1). Acute SFGR was associated with higher recent temperatures (significantly higher nighttime mean (Odds ratio (OR): 1.17 [1.03-1.34], p-value = 0.01) and nighttime maximum (OR: 1.16 [1.02-1.32], p-value = 0.02) temperature at one-month lag and daytime minimum (OR: 1.08 [1.00-1.17], p-value = 0.03) temperature at one-month lag) as well as lower minimum NDWI (a proxy for plant water stress, where lower values signify increased plant stress) at one- and two-month lags (OR:0.20 [0.05- 0.71], p-value = 0.01; OR: 0.09 [0.02-0.38], p-value < 0.001). Additionally, acute SFGR participants had lower minimum evapotranspiration rates at a two-month lag (OR:0.78 [0.64-0.96] p-value=0.02) and higher maximum evapotranspiration rates at a three-month lag (OR:1.06 [1.01-1.12], p-value=0.03).

### Performance of clinical, climate, and environmental predictors and parsimonious model selection

Table 2 lists the best performing clinical, climate, and environmental predictors, as well as the best performing predictors when combined. We first assessed model performance with only the ten best performing clinical predictors: AUC of 0.64 (95% CI:0.48-0.80) and with only the ten best performing climate and environmental predictors: AUC: 0.61 (95% CI:0.47- 0.77). Next, we fit a model using the ten best performing clinical and the ten best performing climate and environmental predictors and assessed how this model compared to a model with only the ten best performing clinical predictors. By combining clinical, climate and environmental predictors, the AUC improved to 0.72 (95% CI: 0.57-0.86), though this improvement was not statistically significant (median p-value=0.3, 12% of p-values <0.05). A model with a sensitivity of 70%, 80%, and 90% had a specificity of 61%, 51%, and 33% respectively. *Vice versa*, a model with a specificity of 70%, 80%, and 90% had a sensitivity of 62%, 46%, and 29% respectively.

**Table 2.**
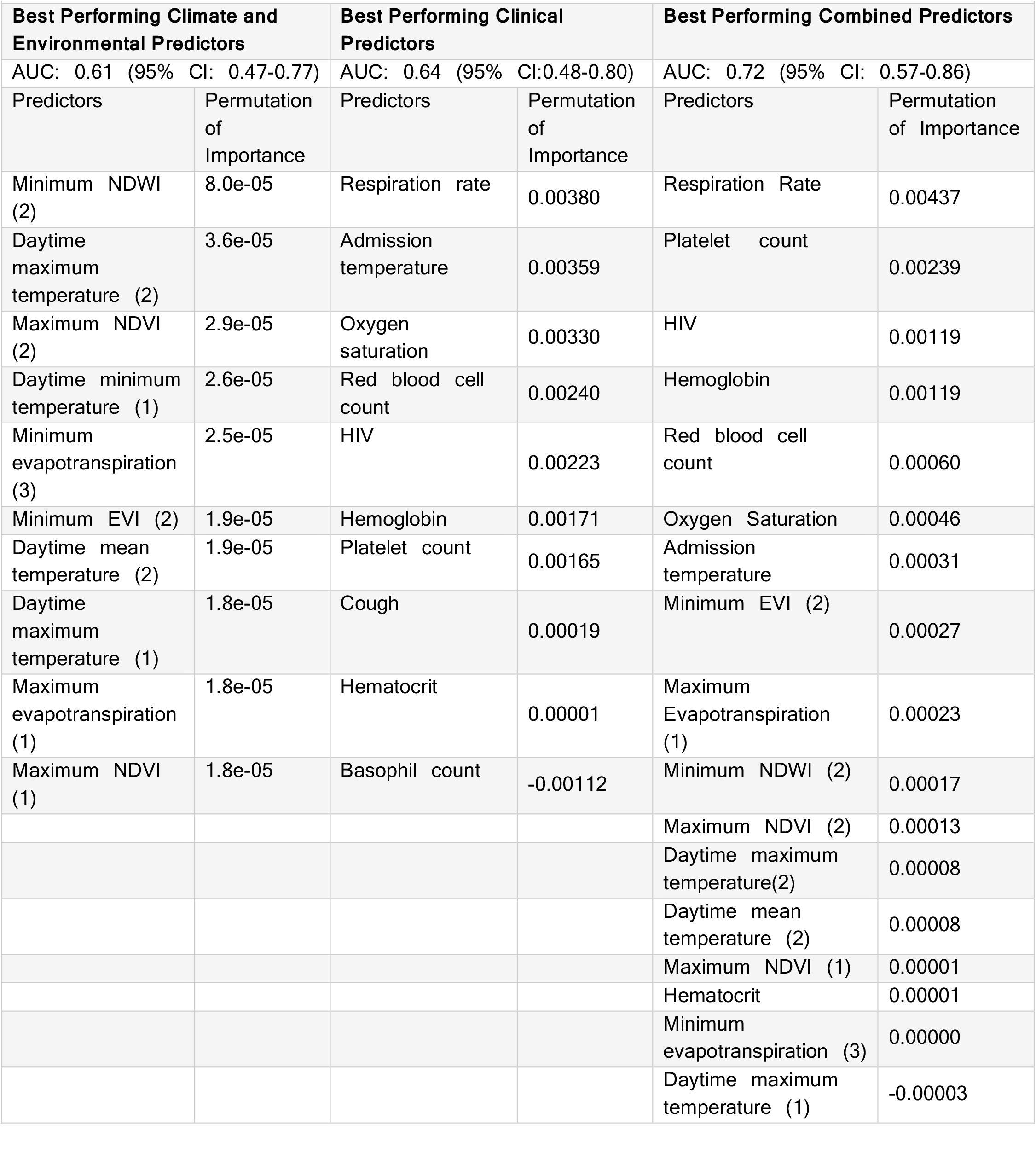
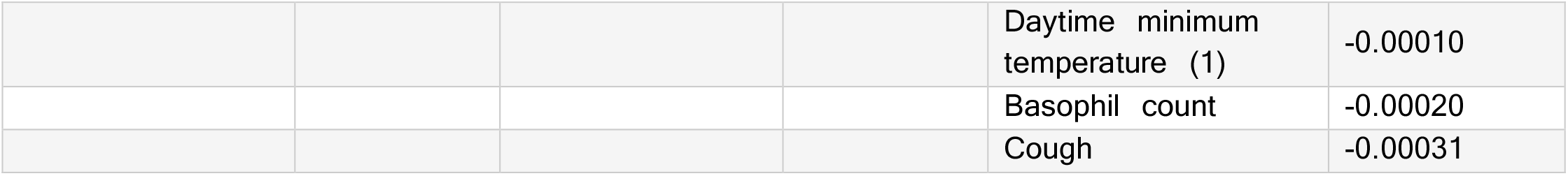
Best performing predictors by permutation of importance for climate, environmental, and clinic Predictors

To create a parsimonious model, we fit models by successively incorporating fewer of the best performing predictors. Model performance was relatively similar with 10, 15, and 20 predictors and began to decrease with less than 10 predictors (Figure 2). A model with the best performing 10 predictors had an AUC: of 0.69 (95%CI 0.54-0.84) and a sensitivity of 66%, a specificity of 72%, PPV of 93%, and NPV of 33%. This model would include eight clinical predictors: respiration rate, platelet count, HIV rapid test result, red blood cell count, hemoglobin, admission temperature, oxygen saturation, basophil count and two climate and environmental predictors: minimum NDWI at a two-month lag and maximum evapotranspiration at a one-month lag.

**Figure 2.**
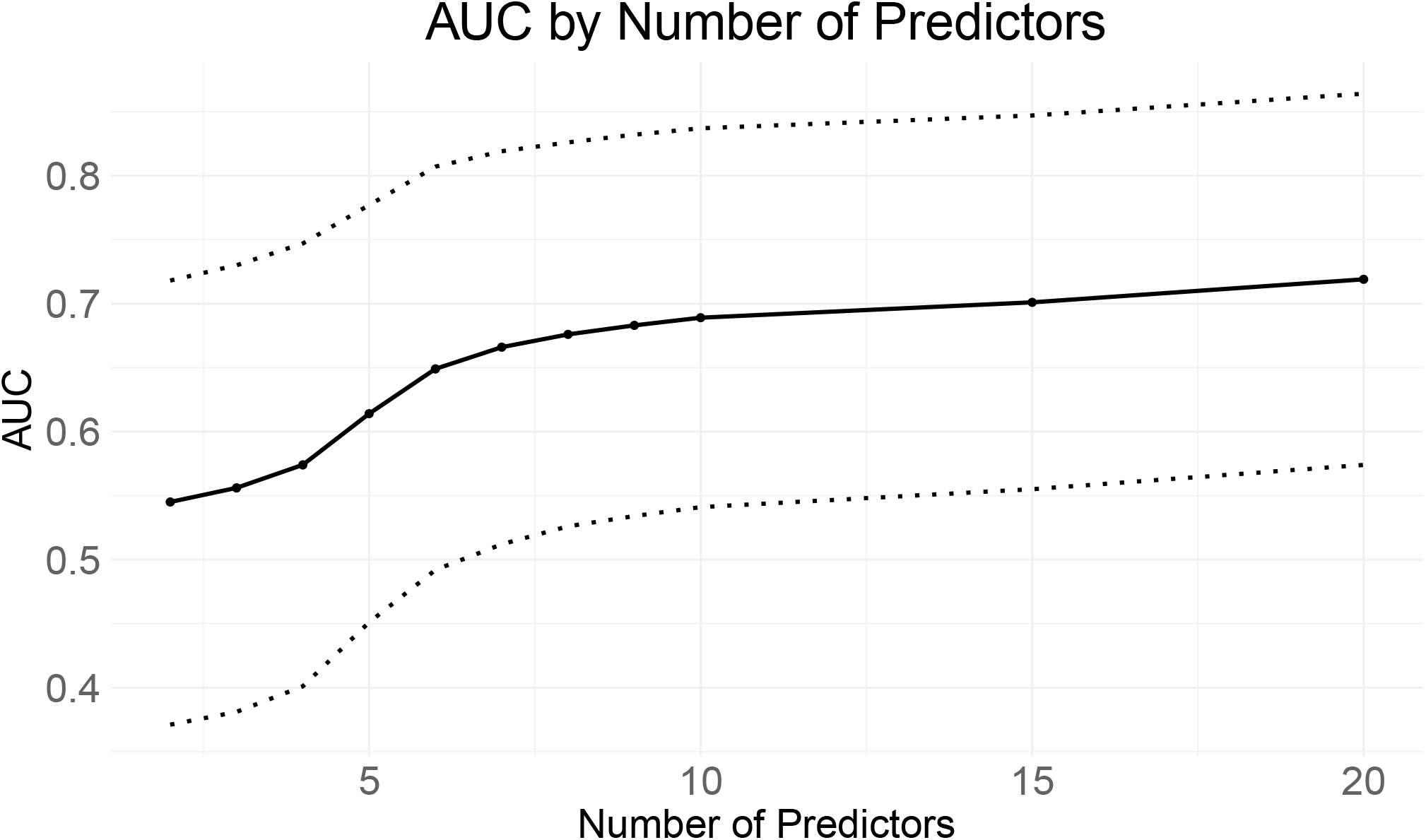
Average AUC (solid line) and 95% Confidence Intervals (dotted lines) from cross-validation (100 iterations) for each model by number of predictors included in the model. AUC: area under the receiver operating characteristic curve.

## Discussion

Using data from a two-center clinical study of febrile illness from northern Tanzania, we show the derivation and cross-validation of a diagnostic prediction model for SFGR, a febrile illness lacking an accurate laboratory diagnostic during acute illness. We also showed that the addition of satellite-derived climate and environmental predictors improved the predictive performance of clinical predictors alone. A parsimonious model with ten predictors including three vital signs, four results from CBC, two satellite-derived climate predictors, and a rapid HIV test achieved an AUC of 0.69 (95%CI 0.54-0.84) on cross-validation. While our predictive model offers an improvement over existing clinical prediction models published for SFGR,^32^ given the suboptimal performance of these models, there is a critical need for the exploration and validation of specific biomarkers that could enhance diagnostic precision of SFGR clinical prediction models and contribute to more effective management strategies in regions affected by this potentially fatal bacterial disease. We propose assessing candidate biomarkers, including proteins, peptides, and nucleic acids, including routine clinical analytes (e.g., fibrinogen) and vetted translational research assays (e.g., endothelial activation markers such as angiopoietein-2) that are relevant to SFGR’s known pathophysiology of endothelial infection and inflammation.

The use of satellite Imagery has been shown to facilitate modeling of population dynamics of ticks,^33,34^ the vector for transmission of SFGR. In our prediction modeling, satellite-derived climate and environmental predictors improved the AUC of our internally validated model. The optimum threshold for the parsimonious model resulted in a sensitivity of 66%, a specificity of 72%, PPV of 93%, and NPV of 33%. A PPV of 93% would allow clinicians to use this model to determine which patients should be started on empiric treatment for SFGR with tetracycline therapy. However, a sensitivity of 66% indicates that this model would miss nearly one-third of SFGR patients. Models with higher sensitivities had much lower specificities (i.e., high potential for false positive predictions). Given the dynamic nature of NDVI, EVI, and NDWI, which undergo variations influenced by land use changes and other anthropogenic impacts,^35–37^ external validation of the model is needed, as these fluctuations may impose limitations on their applicability within models spanning several years. For use in a clinical decision support tool, the most recent satellite-derived climate and environmental data could be gathered from online sources, based on smartphone-based detection of GPS location.

Similar to what has been reported in the literature, we found that a higher body temperature on admission was one of the top clinical predictors associated with acute SFGR infection.^38,39^ CBC results including thrombocytopenia, leukopenia, and lymphopenia have been shown to be significantly different between acute SFGR and non-SFGR febrile illness. In our model, platelet count, hemoglobin, and hematocrit were important CBC predictors, however, lymphopenia and leukopenia were not. Our model also found that respiration rate and oxygen saturation contribute to discrimination between acute SFGR and non-SFGR febrile illness. While our dataset did not have complete information on classic SFGR symptoms including headache, myalgias, and rash,^11^ these symptoms have been found to occur in similar proportions among acute SFGR and non-SFGR febrile illness.^9,10,40^

A major limitation of our study is lack of external validation. The lack of external validation, coupled with the fact that we constructed our model using data from a single endemic Region in northern Tanzania, potentially hinders the model’s generalizability to a broader population. Given the intricate interplay between vector and host, the climate and environmental indices that affect SFGR may vary between regions. Second, our model was constructed using a relatively small dataset, resulting in wide confidence intervals for the calculated cross-validated AUC. could help Finally, our model lacks other laboratory values that have shown to be correlated with SFGR infection (e.g., sodium,^38,41^ transaminases,^38,41–43^ lactic dehydrogenase,^41,43^ fibrinogen^44^); inclusion of these laboratory parameters may have improved model performance.

As a proof of concept using an existing dataset of acute SFGR and non- SFGR febrile illness in Tanzania, we demonstrated proof of concept that inclusion of climate and environmental variables along with clinical variables improved clinical prediction models for identifying SFGR. Further research should expand upon this analysis by incorporating data from additional febrile cohorts, exploring the inclusion of clinical biomarkers, and assessing the performance of this model in diverse settings endemic for SFGR to ensure its generalizability.

## Data Availability

All data produced in the present study are available upon reasonable request to the authors

## Acknowledgements

This research was supported by the International Studies on AIDS Associated Co-infections, United States National Institutes of Health (U01 AI062563 to J.A.C. and V.P.M, K24 AI166087 to D.T.L., and R38 HL143605 to R.J.W. through Utah Stimulating Access to Research in Residency (StARR)). We acknowledge the Hubert-Yeargan Center for Global Health at Duke University for critical infrastructure support for the Kilimanjaro Christian Medical Centre-Duke University

Collaboration. **Disclaimers:** The findings and conclusions in this report are those of the authors and do not necessarily represent the official position of the US Centers for Disease Control and Prevention. Use of trade names and commercial sources is for identification only and does not imply endorsement by the US Department of Health and Human Services or the US Centers for Disease Control and Prevention.

---

**Supplemental Table S1.** TRIPOD checklist.

| Section/Topic | Item | Checklist Item | Page |
| --- | --- | --- | --- |
| **Title and abstract** |  |  |  |
| Title | 1 | Identify the study as developing and/or validating a multivariable prediction model, the target population, and the outcome to be predicted. | 1 |
| Abstract | 2 | Provide a summary of objectives, study design, setting, participants, sample size, predictors, outcome, statistical analysis, results, and conclusions. | 2 |
| **Introduction** |  |  |  |
| Background and objectives | 3a | Explain the medical context (including whether diagnostic or prognostic) and rationale for developing or validating the multivariable prediction model, including references to existing models. | 2-3 |
|  | 3b | Specify the objectives, including whether the study describes the development or validation of the model or both. | 3 |
| **Methods** |  |  |  |
| Source of data | 4a | Describe the study design or source of data (e.g., randomized trial, cohort, or registry data), separately for the development and validation data sets, if applicable. | 3-4 |
|  | 4b | Specify the key study dates, including start of accrual; end of accrual; and, if applicable, end of follow-up. | 3-4 |
| Participants | 5a | Specify key elements of the study setting (e.g., primary care, secondary care, general population) including number and location of centres. | 3-4 |
|  | 5b | Describe eligibility criteria for participants. | 3-4 |
|  | 5c | Give details of treatments received, if relevant. | N/A |
| Outcome | 6a | Clearly define the outcome that is predicted by the prediction model, including how and when assessed. | 4 |
|  | 6b | Report any actions to blind assessment of the outcome to be predicted. | N/A |
| Predictors | 7a | Clearly define all predictors used in developing or validating the multivariable prediction model, including how and when they were measured. | 4 |
|  | 7b | Report any actions to blind assessment of predictors for the outcome and other predictors. | N/A |
| Sample size | 8 | Explain how the study size was arrived at. | 7 |
| Missing data | 9 | Describe how missing data were handled (e.g., complete-case analysis, single imputation, multiple imputation) with details of any imputation method. | 5 |
| Statistical analysis methods | 10a | Describe how predictors were handled in the analyses. | 5 |
|  | 10b | Specify type of model, all model-building procedures (including any predictor selection), and method for internal validation. | 5 |
|  | 10d | Specify all measures used to assess model performance and, if relevant, to compare multiple models. | 5 |
| Risk groups | 11 | Provide details on how risk groups were created, if done. | N/A |
| **Results** |  |  |  |
| Participants | 13a | Describe the flow of participants through the study, including the number of participants with and without the outcome and, if applicable, a summary of the follow-up time. A diagram may be helpful. | 7 |
|  | 13b | Describe the characteristics of the participants (basic demographics, clinical features, available predictors), including the number of participants with missing data for predictors and outcome. | 6-8 |
| Model development | 14a | Specify the number of participants and outcome events in each analysis. | 6-8 |
|  | 14b | If done, report the unadjusted association between each candidate predictor and outcome. | N/A |
| Model specification | 15a | Present the full prediction model to allow predictions for individuals (i.e., all regression coefficients, and model intercept or baseline survival at a given time point). | 9-10 |
|  | 15b | Explain how to use the prediction model. | 9-11 |
| Model performance | 16 | Report performance measures (with CIs) for the prediction model. | 9-10 |
| **Discussion** |  |  |  |
| Limitations | 18 | Discuss any limitations of the study (such as nonrepresentative sample, few events per predictor, missing data). | 11-12 |
| Interpretation | 19b | Give an overall interpretation of the results, considering objectives, limitations, and results from similar studies, and other relevant evidence. | 11-12 |
| Implications | 20 | Discuss the potential clinical use of the model and implications for future research. | 11-12 |
| **Other information** |  |  |  |
| Supplementary information | 21 | Provide information about the availability of supplementary resources, such as study protocol, Web calculator, and data sets. | 12-14 |
| Funding | 22 | Give the source of funding and the role of the funders for the present study. | 12 |

**Supplemental Table S2.** MODIS satellite-derived climate and environmental predictors for acute SFGR and non-SFGR febrile illness, Kilimanjaro region, Tanzania 2007-2008

|  |  |  |  |  |
| --- | --- | --- | --- | --- |
| **Satellite predictor (months lagged)** | **Overall, N=449<sup>1</sup>** | **Non-SFGR febrile illness, N=378<sup>1</sup>** | **Acute SFGR, N=71<sup>1</sup>** | **p-value<sup>2</sup>** |
| **Mean EVI (1)** | 0.28 (0.22, 0.30) | 0.28 (0.23, 0.30) | 0.28 (0.22, 0.29) | 0.2 |
| **Mean NDVI (1)** | 0.47 (0.42, 0.52) | 0.48 (0.42, 0.53) | 0.47 (0.42, 0.50) | 0.5 |
| **Maximum EVI (1)** | 0.71 (0.65, 0.73) | 0.71 (0.65, 0.73) | 0.71 (0.69, 0.72) | >0.9 |
| **Maximum NDVI (1)** | 0.927 (0.923, 0.937) | 0.927 (0.923, 0.941) | 0.927 (0.923, 0.933) | 0.9 |
| **Minimum EVI (1)** | -0.16 (-0.19, -0.12) | -0.16 (-0.19, -0.12) | -0.16 (-0.18, -0.12) | 0.073 |
| **Minimum NDVI (1)** | -0.167 (-0.188, -0.158) | -0.167 (-0.188, -0.149) | -0.167 (-0.186, -0.158) | >0.9 |
| **Mean NDWI (1)** | 0.22 (0.21, 0.30) | 0.23 (0.20, 0.30) | 0.22 (0.21, 0.24) | 0.4 |
| **Minimum NDWI (1)** | 1.11 (1.01, 1.29) | 1.12 (1.03, 1.29) | 1.10 (0.93, 1.14) | 0.008 |
| **Maximum NDWI (1)** | 0.25 (0.22, 0.29) | 0.25 (0.22, 0.29) | 0.24 (0.22, 0.26) | 0.087 |
| **Mean evapotranspiration (1)** | 13.7 (11.1, 17.5) | 13.7 (11.1, 17.5) | 12.9 (12.0, 14.6) | 0.2 |
| **Minimum evapotranspiration (1)** | 3.95 (3.25, 4.60) | 3.95 (3.25, 5.00) | 3.90 (3.25, 4.60) | 0.6 |
| **Maximum evapotranspiration (1)** | 61 (56, 63) | 61 (58, 63) | 61 (55, 64) | 0.4 |
| **Nighttime Mean Temp. (1)** | 288.04 K (286.27, 289.02) | 288.04 K (286.27, 288.93) | 288.86 K (287.42, 290.59) | 0.008 |
| **Nighttime Maximum Temp. (1)** | 295.25 K (293.53, 295.86) | 294.95 K (293.53, 295.86) | 295.76 K (294.34, 297.77) | 0.005 |
| **Nighttime Minimum Temp. (1)** | 266.7 K (265.0, 269.2) | 266.4 K (265.0, 269.2) | 266.8 K (264.9, 269.2) | 0.4 |
| **Daytime Mean Temp. (1)** | 304.1 K (298.7, 306.3) | 303.9 K (298.7, 306.4) | 304.1 K (298.8, 306.3) | 0.14 |
| **Daytime Maximum Temp. (1)** | 317.8 K<br>(309.8, 321.1) | 317.7 K (309.8, 321.1) | 318.1 K<br>(310.5, 321.0) | 0.4 |
| **Daytime Minimum Temp. (1)** | 275.1 K<br>(273.7, 278.6) | 275.1 K (273.7, 278.3) | 276.5 K<br>(275.1, 279.5) | 0.023 |
| **Mean EVI (2)** | 0.28 (0.22, 0.29) | 0.28 (0.22, 0.29) | 0.26 (0.23, 0.28) | 0.2 |
| **Mean NDVI (2)** | 0.47 (0.43, 0.50) | 0.47 (0.43, 0.50) | 0.45 (0.43, 0.50) | 0.5 |
| **Maximum EVI (2)** | 0.71 (0.69, 0.74) | 0.71 (0.69, 0.74) | 0.72 (0.69, 0.74) | 0.2 |
| **Maximum NDVI (2)** | 0.927 (0.923, 0.936) | 0.927 (0.923, 0.936) | 0.928 (0.923, 0.933) | 0.7 |
| **Minimum EVI (2)** | -0.16 (-0.18, -0.12) | -0.16 (-0.18, -0.12) | -0.15 (-0.17, -0.10) | 0.10 |
| **Minimum NDVI (2)** | -0.167 (-0.188, -0.158) | -0.167 (-0.188, -0.158) | -0.164 (-0.186, -0.149) | 0.3 |
| **Mean NDWI (2)** | 0.22 (0.18, 0.24) | 0.22 (0.15, 0.25) | 0.22 (0.21, 0.23) | >0.9 |
| **Minimum NDWI (2)** | 1.12 (1.03, 1.22) | 1.12 (1.05, 1.29) | 1.12 (1.01, 1.14) | 0.011 |
| **Maximum NDWI (2)** | 0.25 (0.22, 0.27) | 0.25 (0.22, 0.27) | 0.24 (0.22, 0.26) | 0.7 |
| **Mean evapotranspiration (2)** | 13.22 (11.47, 15.43) | 13.71 (11.47, 15.43) | 13.22 (12.88, 14.55) | 0.5 |
| **Minimum evapotranspiration (2)** | 3.90 (3.25, 4.55) | 3.90 (3.25, 4.55) | 3.70 (3.25, 4.15) | 0.038 |
| **Maximum evapotranspiration (2)** | 61.1 (58.0, 63.1) | 61.1 (58.2, 63.1) | 60.0 (55.4, 63.1) | 0.3 |
| **Nighttime Mean Temp. (2)** | 288.54 K<br>(287.23, 288.93) | 288.04 K<br>(287.23, 288.93) | 288.54 K<br>(287.36, 289.05) | 0.3 |
| **Nighttime Maximum Temp. (2)** | 295.25 K<br>(293.53, 296.41) | 295.25 K<br>(293.53, 295.86) | 295.53 K<br>(293.57, 297.30) | 0.2 |
| **Nighttime Minimum Temp. (2)** | 266.8 K<br>(265.1, 270.0) | 266.8 K (265.1, 270.0) | 266.8 K<br>(264.5, 270.0) | 0.6 |
| **Daytime Mean Temp. (2)** | 304.1 K<br>(299.8, 306.7) | 304.1 K (299.8, 306.7) | 304.9 K<br>(303.6, 306.7) | 0.2 |
| **Daytime Maximum Temp. (2)** | 318.1 K<br>(310.5, 321.1) | 318.1 K (310.5, 321.1) | 320.0 K<br>(317.6, 321.1) | 0.15 |
| **Daytime Minimum Temp. (2)** | 275.6 K<br>(273.7, 278.6) | 275.4 K (273.7, 278.6) | 275.6 K<br>(273.6, 278.3) | 0.5 |
| **Mean EVI (3)** | 0.26 (0.23, 0.29) | 0.26 (0.22, 0.29) | 0.25 (0.24, 0.28) | 0.5 |
| **Mean NDVI (3)** | 0.45 (0.41, 0.49) | 0.45 (0.41, 0.49) | 0.45 (0.44, 0.52) | 0.11 |
| **Maximum EVI (3)** | 0.71 (0.69, 0.73) | 0.71 (0.65, 0.73) | 0.71 (0.69, 0.74) | 0.5 |
| **Maximum NDVI (3)** | 0.927 (0.923, 0.936) | 0.927 (0.923, 0.936) | 0.927 (0.918, 0.936) | 0.3 |
| **Minimum EVI (3)** | -0.16 (-0.18, -0.12) | -0.16 (-0.18, -0.12) | -0.16 (-0.18, -0.12) | >0.9 |
| **Minimum NDVI (3)** | -0.169 (-0.188, -0.158) | -0.169 (-0.188, -0.158) | -0.183 (-0.188, -0.158) | 0.5 |
| **Mean NDWI (3)** | 0.22 (0.15, 0.23) | 0.22 (0.15, 0.23) | 0.22 (0.15, 0.23) | >0.9 |
| **Minimum NDWI (3)** | 1.12 (1.05, 1.26) | 1.11 (1.05, 1.26) | 1.13 (1.06, 1.29) | 0.15 |
| **Maximum NDWI (3)** | 0.24 (0.22, 0.27) | 0.24 (0.22, 0.27) | 0.24 (0.22, 0.28) | 0.2 |
| **Mean evapotranspiration (3)** | 12.93 (11.42, 14.55) | 12.88 (11.36, 14.55) | 13.71 (11.44, 15.43) | 0.15 |
| **Minimum evapotranspiration (3)** | 3.90 (3.25, 4.35) | 3.90 (3.25, 4.35) | 4.00 (3.25, 4.55) | 0.8 |
| **Maximum evapotranspiration (3)** | 61.1 (57.2, 63.7) | 61.1 (56.4, 63.1) | 62.4 (58.0, 67.4) | 0.024 |
| **Nighttime Mean Temp. (3)** | 288.72 K<br>(287.23, 289.16) | 288.54 K<br>(287.23, 289.16) | 288.72 K<br>(287.35, 288.97) | >0.9 |
| **Nighttime Maximum Temp. (3)** | 295.53 K<br>(293.57, 296.41) | 295.53 K<br>(293.57, 296.41) | 295.74 K<br>(293.66, 296.41) | 0.4 |
| **Nighttime Minimum Temp. (3)** | 266.8 K<br>(265.4, 269.2) | 266.8 K (265.5, 269.2) | 266.7 K<br>(265.1, 269.2) | 0.5 |
| **Daytime Mean Temp. (3)** | 304.89 K<br>(299.81, 306.66) | 304.59 K<br>(299.81, 306.66) | 306.14 K<br>(300.47, 307.02) | 0.10 |
| **Daytime Maximum Temp. (3)** | 319.3 K<br>(311.0, 321.4) | 318.7 K (311.0, 321.4) | 320.3 K<br>(311.7, 321.4) | 0.14 |
| **Daytime Minimum Temp. (3)** | 276.8 K<br>(274.0, 278.7) | 276.8 K (273.7,<br>278.7) | 276.6 K<br>(274.0, 278.7) | 0.8 |
| <sup>1</sup>**Median (IQR)** |  |  |  |  |
| <sup>2</sup>**Wilcox rank sum test** |  |  |  |  |

## Notes

### Competing Interest Statement

The authors have declared no competing interest.

